# Common tandem repeat variants associated with glaucoma risk in individuals of African ancestry

**DOI:** 10.1101/2025.02.19.25322489

**Authors:** Kenneth Pham, Roy Lee, Isabel Di Rosa, Rebecca Salowe, Fangming Jin, Yuki Bradford, Gui-Shuang Ying, Jennifer E. Phillips-Cremins, Shefali Setia-Verma, Joan O’Brien

## Abstract

The contribution of common tandem repeats (TR) variants to common, complex disease remains unknown, especially in populations historically underrepresented in genetic research. We identified common TR variants associated with risk of primary open-angle glaucoma (POAG) in individuals of African ancestry. The POAG-associated TR variants were predominantly found at Alu poly(A) tail elements, regions, retinal development enhancers, and harbor binding sites of a POAG-associated transcription factor, LMX1B, suggesting a convergent mechanism of how common TR variation arises and contributes to POAG pathophysiology.

## Main

Glaucoma is the leading cause of irreversible blindness worldwide^1^. Primary open-angle glaucoma (POAG), the most common form of the disease^2–4^, disproportionally affects populations of African ancestry^3^ with more severe disease outcomes^5^. Since early stages of POAG are asymptomatic^6^, with approximately half of patients unaware they have the disease^7^, tools that identify individuals at risk of developing POAG are needed to enable earlier intervention to prevent disease progression and preserve vision.

Several large genome-wide association studies (GWAS) in multi-ancestry populations^8–10^ and individuals of African ancestry^11^ have identified POAG risk-associated loci. Primarily, studies followed up with these risk-associated SNPs by implicating nearby genes and the biological pathways in which these genes act^8,10,11^. However, genes constitute a minority of the genome, and other genetic elements, such as tandem repeats (TRs), contribute to diversity across human genomes. A recent GWAS found an association between a variable tandem number repeat (VNTR) in *TMCO1* and POAG in individuals of European ancestry^12^. Here, we investigated whether common TR variants associate with POAG risk in individuals of African ancestry.

First, we leveraged four large-scale GWAS conducted in African ancestry or multi-ancestry populations: (1) POAG mega-analysis in three cohorts of African ancestry individuals, including the Primary Open-Angle African American Glaucoma Genetics (POAAGG) study^11^; (2) multi-ancestry POAG GWAS^10^; (3) multi-ancestry POAG GWAS across 15 biobanks globally^8^; and (4) a multi-ancestry POAG GWAS with the Department of Veterans Affairs Million Veteran Program (MVP) data^9^ (**Fig 1a**). With the compiled genome-wide significant SNPs, we identified SNPs in high linkage disequilibrium with the POAG-associated SNPs in individuals of African ancestry. We then leveraged a reference panel of phased TRs and SNPs^13^ built with whole genome sequencing (WGS) data from individuals of African ancestry (**Methods**). We filtered for TRs with unit repeat length greater than two, as reported disease-associated TRs have predominantly been repeat unit lengths of three or greater^14^, yielding 17 potential TRs (**Table 1**).

**Fig 1.**
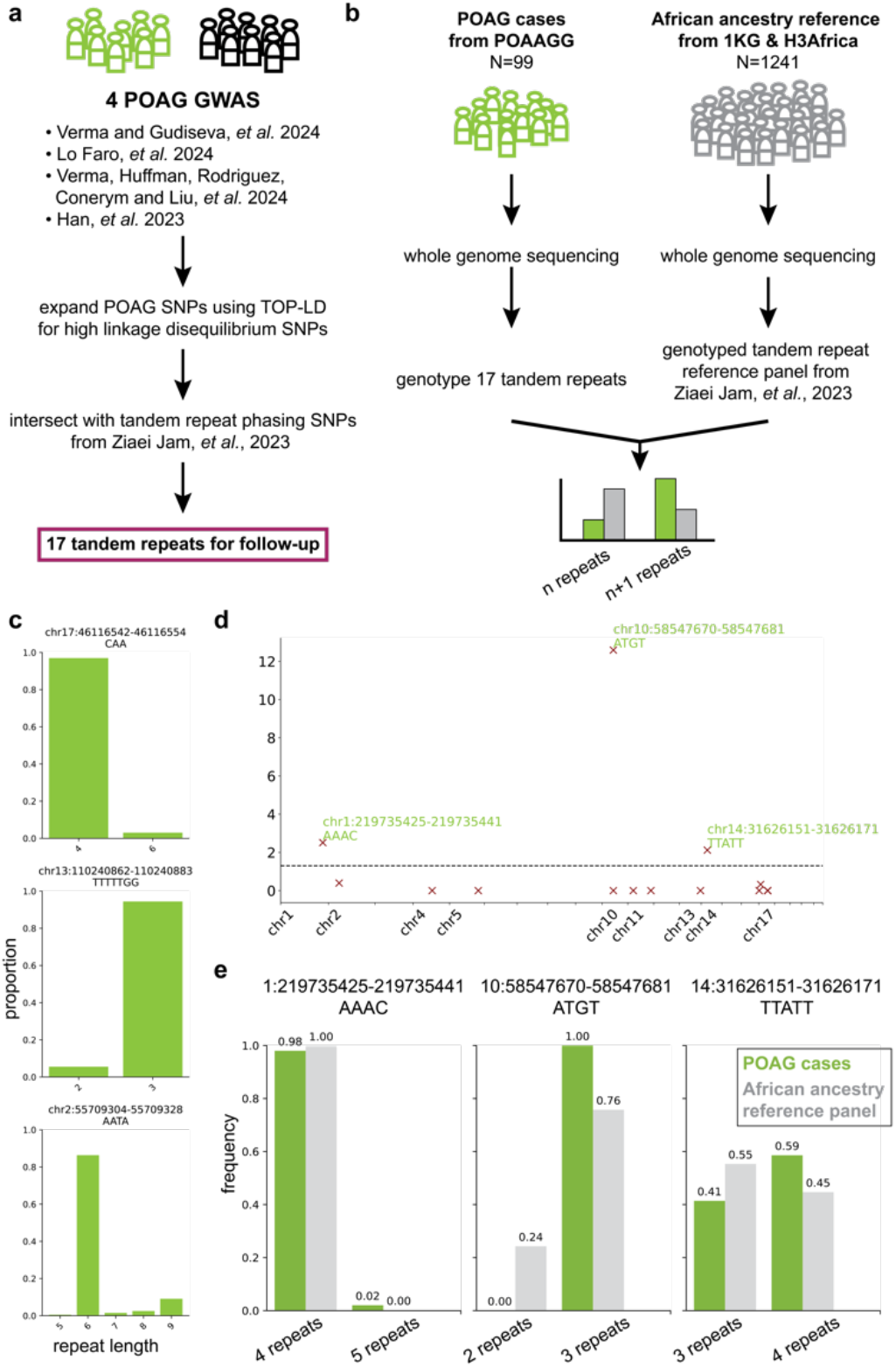
Tandem repeat variants associated with POAG risk in individuals of African ancestry. **a**, Schematic summarizing approach to nominate tandem repeats, starting with 4 large GWAS conducted in populations of multiple ancestries^8–10^ or individuals of African descent^11^, identifying SNPs in high linkage disequilibrium, and intersecting with SNPs that phase tandem repeats in individuals of African ancestry^13^. **b**, Schematic of approach to identify tandem repeat variants associated with POAG risk in individuals of African ancestry. **c**, Three tandem repeat length distributions from WGS of POAG cases in the Primary Open-Angle African American Glaucoma Genetics (POAAGG) study. **d**, Manhattan plot of 17 nominated TRs from **a** and their association with POAG in individuals of African ancestry. **e**, Frequency of TR lengths in POAG cases from the POAAGG study and the African ancestry reference panel^13^ for the 3 significant TRs associated with POAG identified in **d**.

To test if any of the nominated TRs were associated with POAG risk, we conducted WGS on 99 individuals of African ancestry with POAG in the POAAGG study^15^ (**Fig 1b**). We found that within the POAAGG WGS dataset, 16 out of 17 of the nominated TRs were polymorphic (**Fig 1c, Suppl. Fig. 1**), with 11 being dimorphic, and 5 with greater than two alleles in our study population (three allele lengths: 3 TRs; four allele lengths: 1 TR; and five allele lengths: 1 TR). When compared to a reference dataset of TR from an ancestry-matched population^13^ (**Fig 1b, Methods**), we found that 3 of the 17 TRs (chr1:219735425-219735441 AAAC; chr10:58547670-58547681 ATGT; chr14:31626151-31626171 TTATT, hg38) had a differential proportion of allele lengths in the POAG cases relative to the ancestry-matched population comparison (FDR = 5%) (**Fig 1d, Suppl. Fig. 2a**).

**Fig 2.**
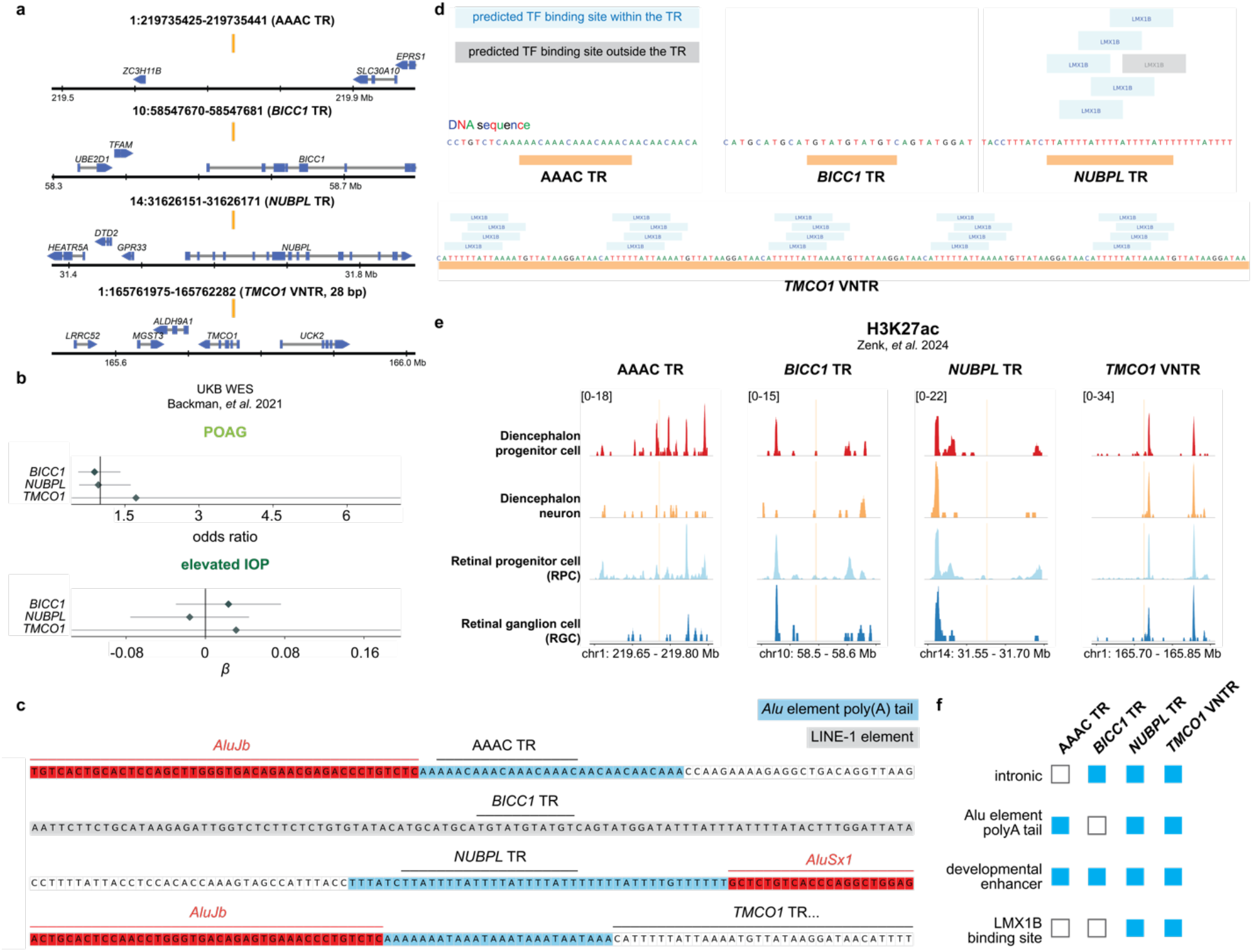
POAG risk-associated tandem repeats harbor binding sites of POAG-associated transcription factors and localize at development enhancers at poly(A) tails of *Alu* elements. **a**, Genomic location of the three TRs associated with POAG in individuals of African descent and the VNTR associated with POAG in individuals of European descent from ref.12^12^. **b**, Association of aggregates pLOF and deleterious missense variants with POAG and elevated IOP from ref.17^17^. **c**, Genomic context of the four POAG-associated TRs with repetitive elements labeled. (Alu elements in red, Alu element poly(A) tails in blue, and LINE element in grey.) **d**, Predicted binding sites at the POAG-associated TRs of the POAG-associated TFs, *LMX1B* and *SIX6*. (Binding sites within the TR in blue, and binding sites not contained by the TR in grey.) **e**, H3K27ac single nucleus CUT&Tag signal across retinal development from human retinal organoids, psuedo-bulked by cell type^24^. **f**, Summary of shared characteristics of the four POAG-associated TRs.

Since an intronic VNTR in *TMCO1* has recently been associated with POAG risk in individuals of European ancestry^12^, we compared the frequency of expanded intronic *TMCO1* TR in POAG cases to the frequency in individuals of African ancestry in the 1000 Genomes Project^12,16^. We found that the expanded *TMCO1* TR had a frequency of 11.6% in the POAG cases compared to 8.3% in the population reference (**Suppl. Fig. 2b**). Together, our data contributes three new common TR variants associated with POAG risk to the previously identified *TMCO1* VNTR variant.

Of the four POAG-associated TR variants, three of the four TRs identified fall into the intron of protein coding genes: the ATGT TR in *BICC1*, the TTATT TR in *NUBPL*, the 28 bp VNTR in *TMCO1* (**Fig. 2a**). To assess whether the protein products of these genes are implicated in contributing to POAG risk, we accessed UK Biobank gene-trait association studies based on whole exome sequencing (WES) data^17^. We assessed *BICC1, NUBPL* and *TMCO1* for associations with POAG or elevated intraocular pressure (IOP) (**Fig. 2b**) and did not find evidence of association with either POAG or elevated IOP.

Rare, penetrant TR variants—especially intronic TRs with AT rich sequences—that result in human disorders often localize at the poly(A) tail of Alu elements^18,19^. We assessed whether the common, POAG-risk associated TR variants share characteristics with rare, penetrant TR variants. Among the TRs associated with POAG, we found that all but one lie within or adjacent to an *Alu* element poly(A) tail (**Fig. 2c**). The AAAC TR and the *TMCO1* TR localize to the poly(A) tail of an *AluJb* element. We found the *NUBPL* TR within the poly(A) tail of an *AluSx1* element. The exception, the *BICC1* TR, lays within a LINE-1 repeat. Similar to rare, penetrant variants^18,19^, POAG-risk associated TR variants predominantly localize to poly(A) tails of *Alu* elements.

Rare, penetrant TR variants can modify disease phenotypes through influence on gene regulation^18,20-22^, including serving as sites of transcription factor (TF) binding^18,23^. To investigate whether the TRs could be TF binding sites of relevance to POAG, we considered TFs with human genetic evidence of association with POAG. Manual curation of the primary literature identified ten genome-wide significant loci where the POAG-associated SNPs are either in a gene or the nearest gene encodes for a TF (**Suppl. Fig. 3**). We utilized the UK Biobank WES data^17^ to assess which of these TFs associated with POAG risk and found *LMX1B* and *SIX6* variants associated with greater risk of POAG (**Suppl. Fig. 3**). When we assessed the four POAG-associated TRs, we found predicted binding sites for *LMX1B* within the TTATT TR in *NUBPL* and in *TMCO1* VNTR, but not within the other two TRs (**Fig. 2d**).

Next, we considered to assess the regulatory potential of these TRs, we analyzed single cell epigenomic data from developing human retinal organoids^24^, specifically H3K27ac, a histone modification that marks enhancers. We found that H3K27ac marks or borders all four TRs across the developmental stages from diencephalon to retinal ganglion cells, with the most pronounced co-localization in retinal progenitor cells (**Fig.2e**).

Together, these data show TRs with POAG-associated risk variants tend to be intronic TRs with binding sites for *LMX1B*, the POAG-associated TF, located at Alu element poly(A) tails with evidence of enhancer activity during retinal development (**Fig. 2f**).

In conclusion, we report TR variants associated with POAG in individuals of African ancestry. Using epigenomic and genomic analyses, we nominated the POAG-associated TRs as putative enhancers in the development of retina and binding sites of a POAG-associated TF, *LMX1B*. Additionally, POAG-associated TR expansions lie within gene introns, specifically adjacent to or within Alu element poly(A) tails. Together, these data inspire a model wherein Alu element poly(A) tails predispose for TR variation as a general hotspot of TR variants. Consequently, TR variants modulate regulatory activity via TF binding to contribute to the pathophysiology of POAG.

**Supplementary Fig. 1.**
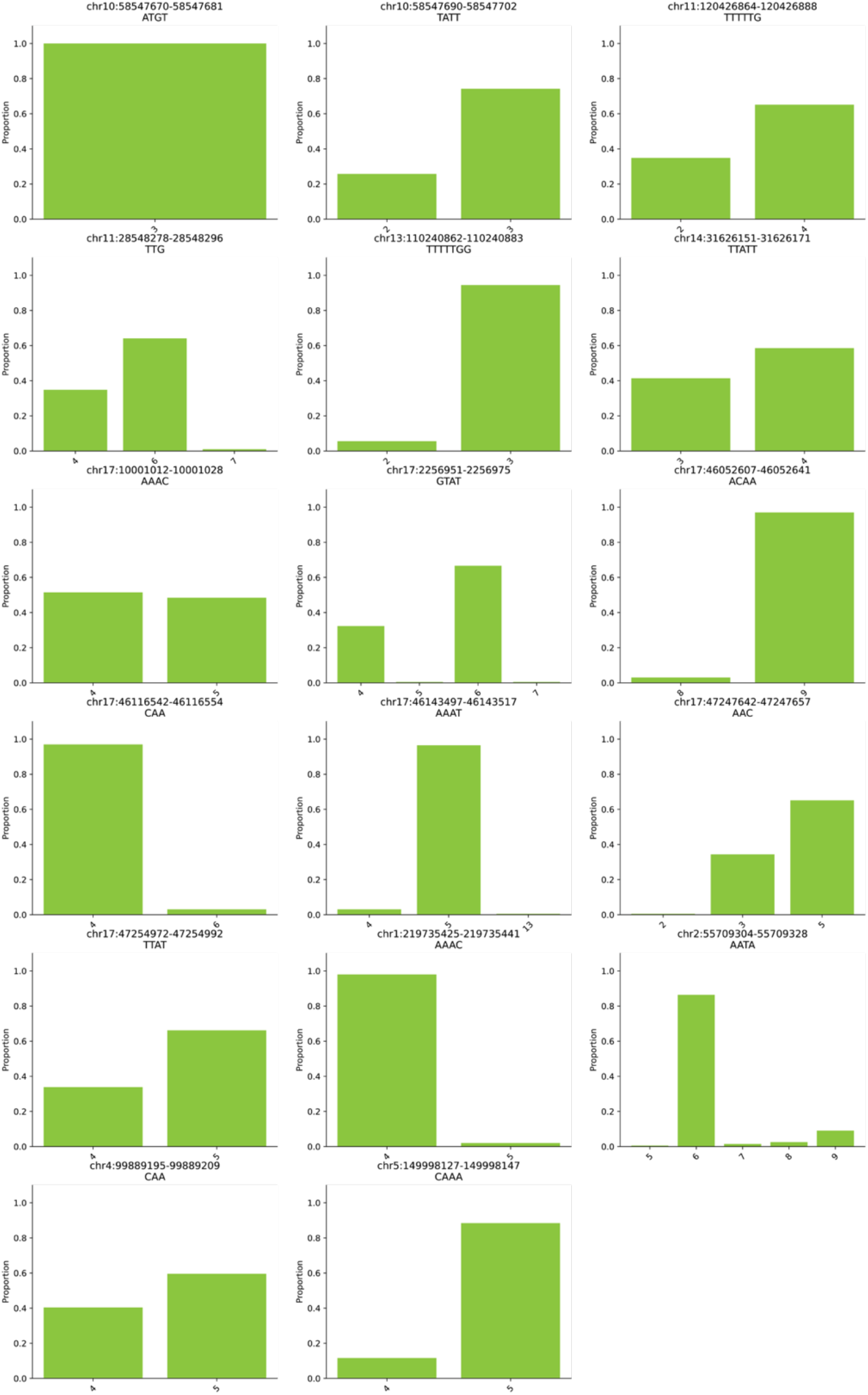
Distribution of tandem repeat lengths in POAG cases. Tandem repeat length distributions from WGS of POAG cases in the POAAGG study for the 17 nominated TRs in **Fig. 1a**

**Supplementary Fig. 2.**
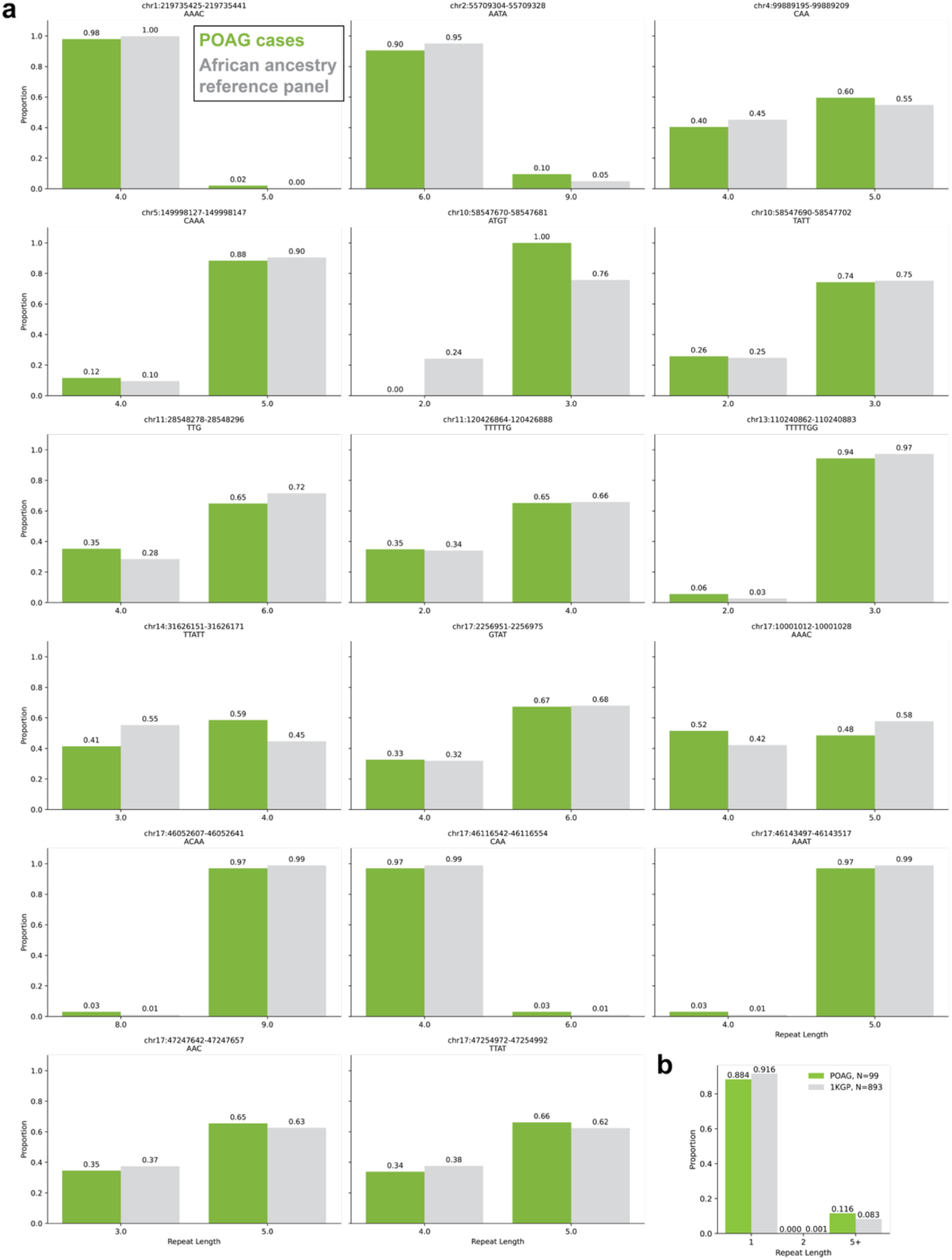
Comparison of tandem repeat lengths in POAG cases and African ancestry reference panel. **a**, Tandem repeat length distributions for the two most common alleles from WGS of POAG cases in the POAAGG study for the 17 nominated TRs compared to the distribution found in an African ancestry reference panel^13^. **b**, Length distributions for a VNTR associated with POAG in individuals of European descent identified in ref.12^12^ in the POAG cases and 1KGP individuals of African ancestry.

**Supplementary Fig. 3.**
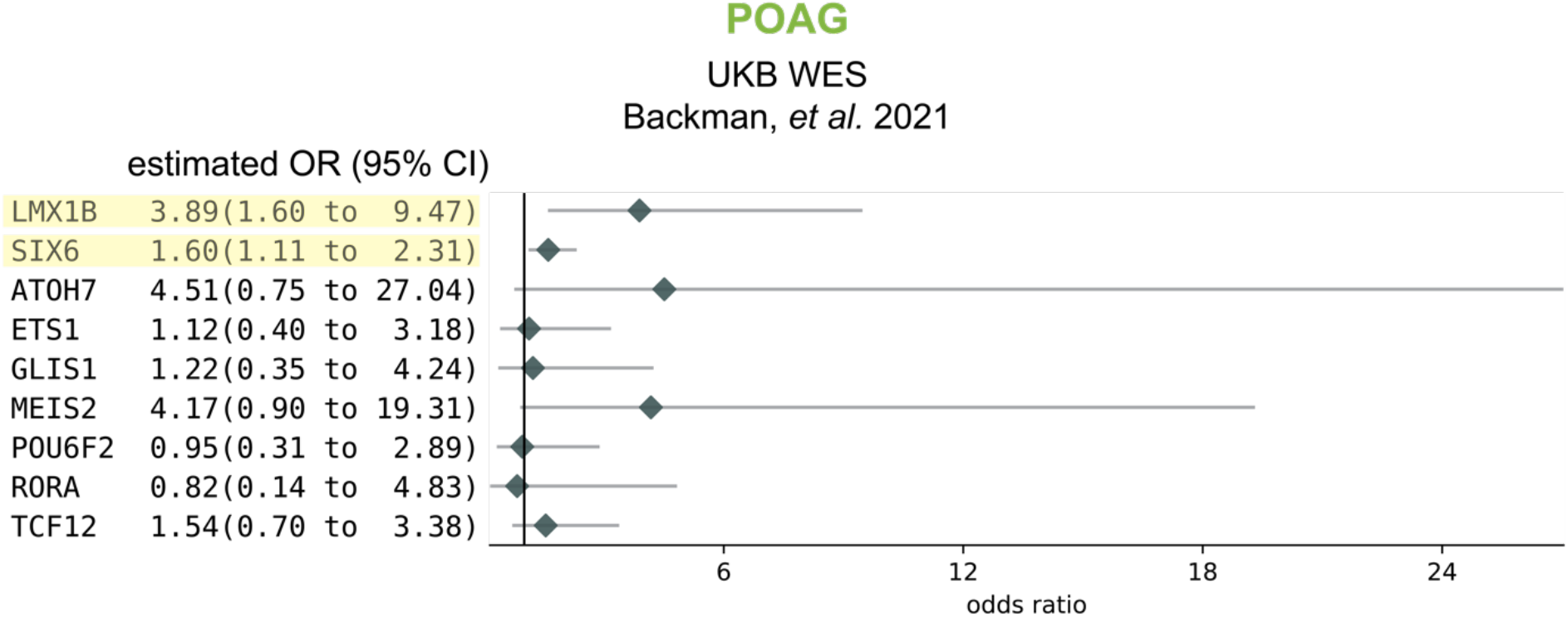
Association of transcription factors with POAG. Association of aggregate pLOF and deleterious missense variants with POAG and elevated IOP from ref.17^17^ for transcription factors with evidence of association with POAG from human genetic studies (*LMX1B*^25,26^, *SIX6*^27^, *ATOH7*^28^, *ETS1*^29^, *GLIS1*^29^, *MEIS2*^25^, *POU6F2*^29^, *RORA*^30^and *TCF12*^29^). Estimated odds ratio and 95% confidence interval (CI) are included.

## Methods

### Identification of TRs associated with POAG

We compiled SNPs associated with POAG from four studies^8–11^. Then, we used TOP-LD^31^ to identify SNPs in high linkage disequilibrium with the POAG-associated SNPs with the following settings: African ancestry; R^2^ > 0.9; MAF > 0. Next, we intersected the expanded list of SNPs a SNP-TR reference panel^51^ to nominate TRs for further study.

### POAAGG study population

The POAAGG study population includes self-identified African ancestry subjects (Black, Afro-Caribbean, or African American), aged 35 years or older, recruited from the Philadelphia area. Glaucoma specialists or ophthalmologists classified subjects as cases, controls, or suspects based on previously published criteria^15^. All subjects signed an informed consent form under IRB-approved protocols. The University of Pennsylvania Institutional Review Board approved the study, which adheres to the tenets of the Declaration of Helsinki.

### Whole genome sequencing

We selected 99 cases from the POAAGG study for WGS. Inclusion criteria for selection included active enrollment at a University of Pennsylvania site (i.e. at least 3 visits over a period of 2 years), without any confounding ocular disease. Of these cases, individuals with a visual acuity of 20/200 or worse, mean deviation on visual field testing of −14dB or worse, and/or average retinal nerve fiber layer thickness less than 60 μm were selected. A manual chart review was performed to ensure that POAG was the primary cause of vision loss. We collected whole blood and isolated DNA. PCR-free WGS was performed at the New York Genome Center to generate 30x coverage whole-genome sequences.

### Risk-associated tandem repeat variant identification

To search for tandem repeat variants associated with POAG risk in African ancestry individuals, we used ExpansionHunter^32^ to genotype the 99 POAG cases with WGS because systematic assessment of tandem repeat genotyping tools demonstrated that ExpansionHunter had the highest accuracy^33^. We genotyped the 17 nominated TRs (see **Identification of TRs associated with POAG**). We then compared the proportions of the two most common allele lengths in POAG cases to the proportions in individuals of African ancestry^13^ that used WGS data from over 1200 individuals of African ancestry^13^ from the 1000 Genomes Project^16^ and H3Africa^34^ cohorts. We used a chi-square test with Bonferroni correction and an FDR of 5%. For the *TMCO1* VNTR, we genotyped the 99 POAG cases using code from ref. 12^12^.

### UKB WES for associations with POAG and elevated IOP

We accessed summary statistics for the whole-exome sequencing data from the UK Biobank. We tested associations by using the odds ratio of POAG and/or elevated IOP with burden tests that combined predicted loss-of-function and predicted deleterious missense mutation with minor allele frequency 1%.

### Alu element poly(A) tail annotation

We annotated Alu elements and their poly(A) tails based on RepeatMasker^35^ annotations accessed on the UCSC Genome Browser^36^.

### Predicted transcription factor binding sites

We mined JASPAR^37^ and HOCOMOCO^38^ for DNA binding motifs for the POAG-associated TFs, *LMX1B* and *SIX6*, and then used FIMO^39^ to search for predicted binding sites at the two TRs with POAG-associated variants.

### H3K27ac data from human developing retinal organoids

We accessed H3K27ac single-nucleus CUT&Tag data from ref.24^24^ from 12 weeks old human retinal organoids for four cell types: diencephalon progenitor cells, diencephalon neurons, retinal progenitor cells, and retinal ganglion cells.

## Data availability

The WGS of the POAG cases in the POAAGG study are available from the corresponding author on reasonable request and with permission of the University of Pennsylvania Institutional Review Board. All other data were publicly available.

## Code availability

We used previously published tools, as indicated in the **Methods** to perform the analyses. No custom code was developed or used.

## Acknowledgements

We thank members of the O’Brien lab for helpful discussions.

